# Whole Exome Sequencing (WES) identifies Homozygous mutation in g.37429732G>A in GRHPR gene is responsible for early onset of nephrolithiasis in the population of West Bengal, India

**DOI:** 10.1101/2022.01.22.22269458

**Authors:** Arindam Chatterjee, Kunal Sarkar, Sarbashri Bank, Siddharth Saraf, Dhansagar wakle, Dilip Kumar pal, Santanu Chakraborty, Sudakshina Ghosh, Biswabandhu bankura, Madhusudan Das

## Abstract

Kidney stone disease (KSD) or nephrolithiasis (NL), is a serious clinical concern that gradually poses threat to global health and economy. The frequency of KSD is rapidly increasing and about 12% of the Indian population are suffering from this disease. In this scenario, we studied the incidence and age group stratification of KSD in the eastern part of India. Furthermore, patients with GRHPR mutation were highly predominant. Furthermore, real time quantitative PCR expression and protein expression through immunoblotting along with protein activity & computational analysis were carried out in patients with GRHPR mutation. Our findings revealed that the candidates of the early age group are prone to hyperoxaluria moreover a potent and frequent mutation in the GRHPR gene is widely present in this age group. Further, biochemical tests such as serum creatine, serum urea, serum and urinary calcium were carried out for the patients along with age matched controls. Moreover, the actual underlying cause of nephrolithiasis for lower age groups still remains unclear. The study revealed that mutations were more often found in GRHPR, AGXT, and HOGA1 genes in the study cohort.

## Introduction

Kidney stone disease (KSD) or nephrolithiasis (NL), is a serious clinical concern that gradually poses threat to global health and economy. The incidence of KSD is much ambiguous and shows an exponential growth rate globally. The frequency of KSD is rapidly increasing and about 12% of the Indian population are suffering from this disease^1^. The prevalence is found more in north and eastern regions of India and some parts of Punjab, Haryana, Delhi, Maharashtra, Gujarat, Rajasthan compared to southern India^2^. This disease has a complex etiology comprising both genetic and environmental factors^3^. KSD is found almost in all stages of life. Previous studies ascertained only cases of kidney stones without describing the age of onset^4^. Moreover, the actual underlying cause of nephrolithiasis for lower age groups still remains unclear. Only few studies in the western hemisphere have been conducted so far^5,6^. When compared to adults, a far higher percentage of pediatric patients have underlying illness that favours stone development (e.g., metabolic disorders, urinary tract anomalies, infections.)^6^. As per the literature study the rate of the incidence of child nephrolithiasis is increasing rapidly throughout the world. Moreover, hardly any global statistics data is found that is related to child nephrolithiasis^4^. In this scenario, we studied the incidence and age group stratification of KSD in the eastern part of India.

Genetic analysis is an urgent need to identify and solve many underlying pathophysiology that is not present in the present clinical database. In this perspective, whole exome sequencing (WES) combined with sanger sequencing offers a powerful technique to gene identification in rare recessive illnesses ^7^. At a relatively modest cost, WES allows detection of dozens of additional genes^8^.Here we carried out WES to 18 unrelated nephrolithiasis patients in the age group between 1 to 18 and included patients up to 25 years if they have a previous report of kidney stone^8^. The study revealed that mutations were more often found in GRHPR, AGXT, and HOGA1 genes in the study cohort.

Further, biochemical tests such as serum creatine, serum urea, serum and urinary calcium were carried out for the patients along with age matched controls. Our findings revealed that the candidates of the early age group are prone to hyperoxaluria moreover a potent and frequent mutation in the GRHPR gene is widely present in this age group. Furthermore, patients with GRHPR mutation were highly predominant. Furthermore, real time quantitative PCR expression and protein expression through immunoblotting along with protein activity & computational analysis were carried out in patients with GRHPR mutation.

## MATERIALS AND METHODS

### Background survey and sample collection

Background survey was conducted from the year 2009-2019. The details of the surveyed data were given in **fig. 1a and 1b**. Based on the data collected from the background survey, the format of the sample collection was designed effectively. The gist of the sample collection was placed in (Supplementary **Table 1)**. Samples for this experiment were collected from individuals with age groups up to 20 years and above 20 years. All the samples were taken according to the prescribed format given by Daga et al. (2018)^8^.

**Fig 1a:**
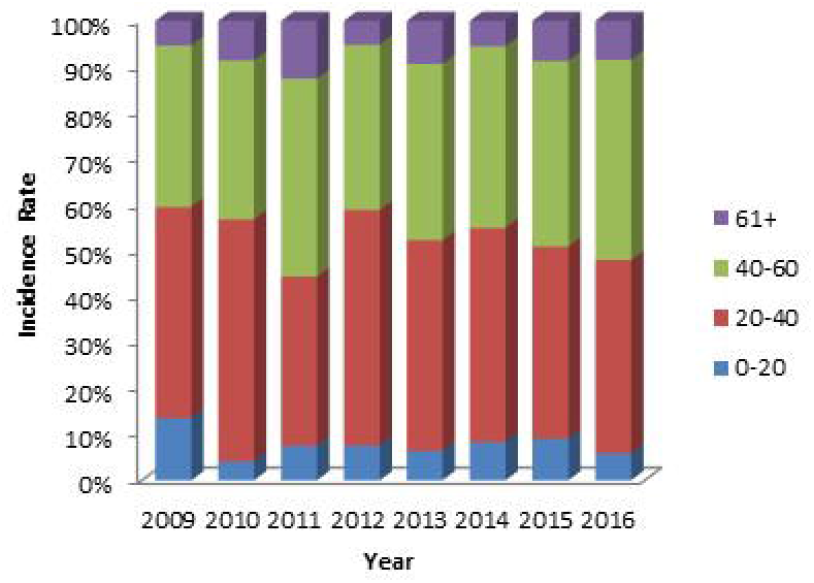

**Fig 1:**
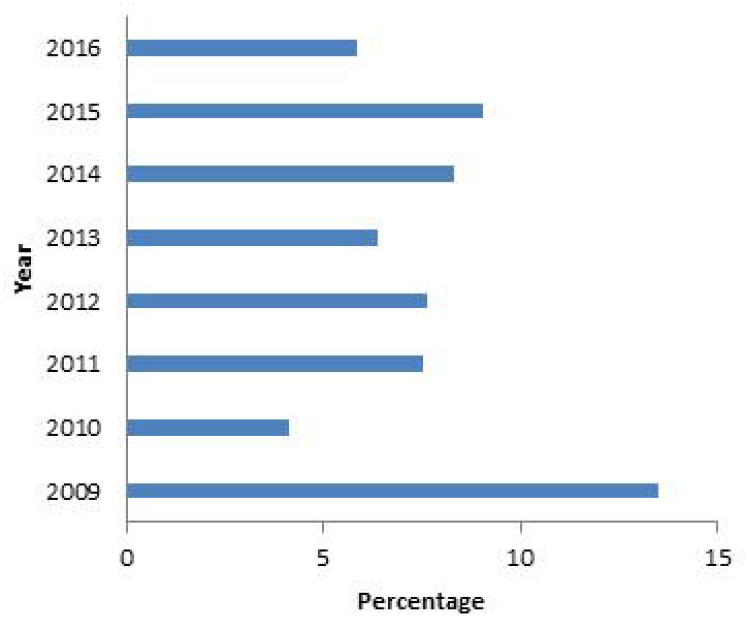

### Study participants

A study was conducted with the participation of patients from the eastern region of India (West Bengal). Patient populations containing kidney stones (s) were identified when renal stones were observed under renal ultrasound, X-ray or multi-detector computed tomography (MDCT). The Ethics Committee of IPGME&R and tertiary care centre of Kolkata, West Bengal India approved the study protocol and informed consent for genetic analysis was obtained from each subject [Ethical no. Inst/IEC/2015/436, dated-07/07/2017]). Sample characteristics of pediatric samples are specified in the Supplementary **table 1**.

### Animal ethical clearance

#### Biochemical analysis

Urine samples were collected and stored in a 5 ml sterile tube containing HCl as a preservative. Creatinine was estimated by using a modified Jaffe’s reaction in an XL-600 analyser (Erba Mannheim, U.S.A.). Serum and urinary calcium were estimated with the application of arsenazo III method in the XL-600 analyser (Erba Mannheim, U.S.A.). Urinary phosphate. Supplementary **table 3**.

### DNA extraction and whole-exome sequencing

Genomic DNA from all participants was isolated from their blood by using DNA QIAamp mini kit [Qiagen] according to the manufacturer’s instructions. Purity of DNA was measured by 260/280 in varioskan lux [Thermofisher]. Whole Exome Sequencing was carried out in the Illumina Hiseq X10 sequencer. Sure selec txt Human All Exon V5+UTR kit [Agilent]was used for library preparation. The generated paired-end fastq files of 150bp were analysed (quality check, trimming, mapping, annoation) using licensed CLC genomics workbench by biomedical genomics plugin. final VCF files were generated and analysed further for variant prioritization.

### Variant prioritization

VCF files were then subjected to many IVA analyses (Qiagen, USA). Similarly, the data were rechecked with the application of various web server mutation distiller^9^, phenolyzer^10^ Moondiploid^11^ Wannovar^12^. Pathogenicity of the found mutation was calculated using Wintervar ^13^following ACMG guidelines.

### Sanger validation & Genotyping

Genomic DNA from samples was amplified along with the mutated region of the GRHPR gene using specific primers listed in **(Table 2)**. The PCR reaction mixture contained 50 ng genomic DNA, 0.5 mM forward and reverse primers, 5% DMSO, and 1x Green master mix [Promega]. The PCR conditions were, Denaturation was done at 95°C for 5 mins followed by 35 cycles of denaturation at 95°C for 30 sec. Annealing was done at 55 °C for 30 sec, extension at 72°C for 30 sec and a final extension at 72°C for 7 min.

### Characterization of kidney stones from samples

Elemental composition of kidney stone samples (n=5) were analyzed using energy disruptive spectroscopy (EDS) [Hitachi S 3400N]. XRD [PANalytical. XPERTPRO] study was done to study the presence of different compounds and crystallinity of the kidney stone samples. The nature of different bonds present in kidney stones and their characteristics was analyzed using FTIR[PerkinElmer spectrum Express Version 1.03.00].

### Bioinformatics

Changes in the genomic sequence in terms of phylop, phastcons Scores were predicted using mutation taster. In order predict the changes in the stability of mutated GRHPR protein stucture in terms of RI and free energy change values (DDG), we used an I-Mutant 2.0 tool that introduced point mutation in that protein and predicted the structural stability. The evolutionary conservancy of amino acid residues of the native GRHPR with that of mutated was determined by ConSurf web server^14^. It also identifies structural residues SNPs of GRHPR protein using evolutionary conservation. Moreover, the Phyre2 homology modeling tool was to produce the 3D structure of native and mutant GRHPR protein. The deleterious SNP was individually replaced in the native sequence of the templates and 3D models for the mutant was predicted Swiss model. **Structural similarities between native and mutant models were investigated based on TM-score and RMSD scores using the TM-align tool**. To substantiate the validity of this finding, we used I-TASSER for further structural analysis of these SNPs in the GRHPR protein. Furthermore, 3D structures of the aforementioned protein were analyzed by SWISS-MODEL to study protein solvation and torsion. The template used for the investigation of the SNPs was 2GCG. Furthermore, mutant protein vs wild protein were subjected to docking using a web server of patchdock..

### Monocyte isolation

Collected fresh blood samples in EDTA coated, or sodium citrate vial and stable it to room temperature. Added Phosphate buffer saline (PBS) into blood in 1:1 ratio in a separate polypropylene tube. In a separate Tube added and histopaque 1.077 in a falcon in 2:1 ratio w.r.t blood PBS. Centrifuge the entire solution in 400g for 35 minutes in 37Cc. A clear phase of monocytes (neutrophils) is observed between both histopaque density and PBMC is seen in the upper phase of histopaque 1.077. Pipetted out the phase and stored in -80°c freezer for further use.

### Real time qpcr [rtqpcr]

Total RNA was isolated from the monocytes of control and patient sample following standard trizol methods. RNA quantity was analysed using varioskan lux multi reader [Thermo Fisher Scientific Inc, USA] by recording absorbance at 260 and 280 nm. Then cDNA was synthesized from patient and control samples using High-Capacity cDNA Reverse Transcription Kit [Thermofisher scientific] according to manufacturer’s protocol. Each qRT-PCR reaction was performed with three biological replicates and three technical replicates. The reaction was performed in 10 μL reaction mixture containing 1 μL cDNA samples as template, 5 μL of 2× SYBR® Green Master Mix (Applied Biosystems, USA), 3 μL of DEPC-treated water and 1 μL of diluted forward and reverse primer mix (10 pmol). The reactions were performed in StepOne™ Real-Time PCR System [Applied Biosystems, USA] Transcript level of all the genes was normalized with an internal reference, *glyceraldehyde 3-phosphate dehydrogenase* (GAPDH) gene from humans. The relative expression ratio of each gene was calculated using the comparative *C*_t_ value method as described in the article^15^. Data represented here are mean values of relative fold change. Values were calculated using ΔΔ*C*_T_ method, and the error bar showed standard deviation. All the primers used in this study are listed in **Table 3**.

### Induced hyperoxaluria in RAT

Male Winstar Rats purchased from NIN, Hyderabad. The rats were maintained under 12 h light and dark cycles at 23°C and 50% relative humidity with free access to food and water ad libitum. All animal procedures were conducted in accordance with the guidelines of Ethical Committee Guidelines [**885/GO/RE/05/CPCSEA DT. 13.4.2017**] for the Care and Use of Laboratory Animals of our institution. A set of 4 groups containing 5 rats in each group were selected based on the number of days of treatment 0,7,14,21 days. Then they were fed 0.75% ethylene glycol with plain drinking water (s.joshi et al, davit T). Oxalate crystals were observed under a bright field microscope in their urine samples from 4th days of treatment showing symptoms of hyperoxaluria(fig 8). Blood samples were collected and monocytes isolated in the respective days for further use.

### Elisa

Samples were incubated with PBS (phosphate buffer) in a 96 well ELISA plate overnight at 4°C, to measure the amount of GRHPR, GAPDH in mice and human samples below. The plates were then rinsed in PBS containing Tween-20 (PBST) buffer before being blocked with 5 mg/ml BSA (bovine serum albumin). Primary antibodies [GRHPR antibody (TA502091), GAPDH ()] were used again after washing the plate. (dilution: 1:200 in PBS) was incubated for 2 hours. Plates were then rinsed three times with PBS-T20 buffer before being treated with 1:10000 goat anti-rabbit IgG (HRP) for 1 hour. After that, the plates were rinsed with PBS-T20 and incubated with the substrate [OPD (1 mg/ml) in 0.05M Citrate-Phosphate buffer. At 450 nm, the colour development was measured and OD values were taken.

### Immunoblotting

Monocytes were isolated from human & rat blood samples Cell lysates were prepared in a RIPA buffer containing protease and phosphatase inhibitors (Sigma) Twenty micrograms(ug) of protein were loaded to each well in SDS-PAGE(10%) and immunoblotting was carried out using standard techniques. Immunoblots were developed using a [Biorad] chemiluminescence method with horseradish peroxidase-labeled secondary antibodies. Densitometric quantification of Western blots was performed by utilizing Image J software (NIH). Protein expression was normalized to loading controls and expressed relative to control conditions. The relative density was calculated in each case (target samples and GAPDH). Then the normalized band intensity of the targets was divided by the normalized GAPDH and the resulting ratio data expressed as Relative density folds change (%), revealing the protein levels and their comparison in the samples.

### Immunocytochemistry

To check the expression of GRHPR Gly165Asp missense mutation in patient’s sample. Isolated monocytes from case and control human samples were placed in 35mm culture dishes and incubated for 3 hours in DMEM (10 percent FBS) for cell attachment. The cells were then rinsed three times with ice cold PBS. The cells are then treated for ICC (ref) using anti-GRHPR antibody and imaged using alexa fluor 555 tagged Anti Mouse secondary antibody in the Floid cell imaging system [Thermofisher]. To identify between the artefacts, the cells were also stained with DAPI. **(Data not provided here)**

### Grhpr assay

The activity of whole proteins extracted from patients’ monocytes was compared to that of controls. Activity measurements were carried in a UV-Vis 660 spectrophotometer in uv range (260nm). Protein activity was measured with PBS buffer (pH 8.0). For the four possible substrate/cofactor combinations, concentrations of glyoxylate with NADPH were used with protein concentrations. Each reading was taken in triplicate. Results are summarized using GraphPad Prism (San Diego California USA, www.graphpad.com) were used for calculation.

kinetic parameters were determined by using the initial velocity (V0) on varying the concentration of the substrate. Concentrations of glyoxylate (0.05–8 mM) and NADH (0.5 mM) or NADPH (0.5 mM). Enzyme concentration was determined by absorbance at 260nm bu measuring NADP. The data were fitted to a Michaelis–Menten equation or substrate inhibition equation to extract the values of the Michaelis constant Km, Vmax and substrate inhibition constant using GraphPad Prism and ms excel.**(Data not provided here)**

## Results

Survey analysis showed that the age group of 0-20 years (<10 %) are less frequent in comparison to the other age groups. The incidence level of the disease was too low but a steady increase over the year (fig 1a & fig1b). Our study assumes a high probability of genetic mutations in the early age of KSD patients since disease prognosis started during early stage in life (craig b langman). We performed WES in 18 individuals of unrelated families with NL and 22 controls. Among the 18 individuals, 16 patients showed genetic mutation causing NL(Table 1). Out of which 8 paediatric patients had pathogenic mutation with hyperoxaluria causing genes with calcium oxalate stones. All affected individuals had hypercalciuria or hyperoxaluria in the presence of NL Only 2 individual among the KSD patients showed no mutation and remained undetected.

**Table 1.** mutation found in all the pediatric samples.

| SL. no | softwares | Gene name | Present in cases | Zygosity | pos(grch37) | rs number | Change in DNA | ref seq | Amino acid change | Novelity | pph2 | pathogenicity | ACMG |
| --- | --- | --- | --- | --- | --- | --- | --- | --- | --- | --- | --- | --- | --- |
| 1 | moon, MD, pheno lyzer | GRHP R | KS1, S4, KS10, KS12, KS14 | HOM(1st reporting homo) | 9:37429729 | rs180177314 | G>A | NM_012203 | c.G494A:/p.G165D | 1st report of homozygous | 1 | pathogenic | PM1,PM2, PP3, PP5,PP6 |
| 2 | do | SERPINH1 | KS1 | HET | 11:75277687 | rs541595707 | G>A | NM_001235 | S98N | no | 0 | benign | PM1,PM2, PP2 |
| 3 | do | AGXT | S4 | comp HET | 2:241808588 | rs180177180 | T>A | NM_000030 | I56N splicing impaired | no | 1 | pathogenic | PM2,PP3,PP5 |
| 4 | do | AGXT | KS4 | HOM(frameshift) | 2:241817038 | rs398122322 | c.33 dup C | NM_000030 | p.lys12fs | yes | NA |  | uncertain significance |
| 5 | do | AGXT | KS6 | HET | 2:241808325 | not assigned | C>T | NM_000030 | L15F | yes | NA | uncertain significance | PM2,BP4 |
| 6 | do | FBN1 | KS6, KS5 | HET(ACMG) | 15:48782174 | rs112287730 | C>T | NM_000138 | A986T | no | 0.458 | possibly damaging | PM2,BP3,BS2,BP1 |
| 7 | do | FBN1 | KS4 | HET(ACMG) | 15:48764757 | not assigned | C>T | NM_000138 | A1443T | yes | 0.316 | likely pathogenic | PM2,B1 |
| 8 | do | HSPG2 | KS1 | HET | 1:22175394 | not assigned | C>A | NM_005529 | G2526V | yes | 0.022 |  | PM2,PP2 |
| 9 | do | HSPG2 | KS5 | HET | 1:22191473 | rs138460117 | A>T | NM_005529 | F1497I | no | 0.997 | damaging | PP2,BS1,BS2 |
| 10 | do | ACVRL1 | KS1 | HET | 12:52307462 | not assigned | C>T | NM_000020 | R145W | yes | 0.001 | uncertain significance | PM1 |
| 11 | do | ATP6<br>V1B1 | S4,K<br>S1 | HET | 2:7119210<br>3 | rs142905<br>621 | G>A | NM_00169<br>2 | R465H |  | 0.997 | probabl<br>y<br>damagin<br>g | PM1,PP3,BS2, BP6 |
| 12 | do | MOCS<br>1 | S4 | HET | 6:3989350<br>5 | rs751538<br>238 | C>T | NM_00594<br>3 | R112Q | no | 0.131 | uncertai<br>n<br>signifiga<br>nce | PM2 |
| 13 | do | CRTA<br>P | KS6 | HET | 3:3315565<br>7 | rs553076<br>085 | C>T | NM_00637<br>1 | R30C | no | 0.999 | probabl<br>y<br>damagin<br>g | PM1,PM2,PP3,BS2,B<br>P1 |
| 14 | do | HOGA<br>1 | KS6 | HET | 10:993616<br>47 | not<br>assigned | T>A | NM_13841<br>3 | V245D | yes | 1 | damagin<br>g | PM1,PM2,PP3,BP1 |
| 15 | do | HOGA<br>1 | S13 | HET | 10:993617<br>47 | rs770050<br>262 | G>A | NM_00113<br>4670 | A115A | no | 0 | benign | PP3,PP5 |
| 16 | do | USP8 | KS7 | HET | 15:507694<br>90 | not<br>assigned | T>C | NM_00515<br>4 | S338P | yes | 0.997 | probabl<br>y<br>damagin<br>g | PM1,PM2 |
| 17 | do | CACN<br>A1D | KS2 | HET | 3:5377708<br>4 | rs567068<br>933 | A>G | NM_00112<br>8840 | N953S | no | 0.666 | possibly<br>damagin<br>g | PM1,PM2,PP3 |
| 18 | do | COL1<br>A2 | KS12 | HET | 7:9404574<br>7 | not<br>assigned | G>A | NM_00008<br>9 | A599T | yes | 0.891 |  | PM1,PM2,BP3,BS1 |
| 19 | do | COL1<br>A2 | KS2,<br>k15 | HET | 7:9403753<br>4 | rs375401<br>215 | G>A | NM_00008<br>9 | A227T | no | 0.181 | benign | PM1,PM2,PP3,BS2,B<br>P6 |
| 20 | do | VIPAS<br>39 | KS3 | HET | 14:779106<br>62 | rs372813<br>446 | C>T | NM_00119<br>3315 | R176H | no | 1 | probabl<br>y<br>damagin<br>g | PM1,PM2,BP1 |
| 21 | do | ATP7<br>B | KS11 | het | 13:525089<br>84 | rs148081<br>616 | C>T | NM00053 | D1229N | no | 0.999 | probabl<br>y<br>damagin<br>g | PM1,PM2 |
| 22 | do | ATP6<br>V1B1 | KS12 | het | 8:2006201<br>9 | rs200124<br>277 | A>G | NM001693 | p.N54S | no | 0.024 | BENIGN | PM1,PM2,BS2 |
| 23 | do | CLIP2 | KS14 | het | 7:7375319 | rs617399 | C>T | NM0038 | P179S | no |  |  | PM1,BP6 |
|  |  |  |  |  | 1 | 91 |  |  |  |  |  |  |  |
| 24 | do | VDR | KS15 | het | 12:482589<br>47 | not<br>assigned | G>A | NM000376 | R54W | YES | 1 | probabl<br>y<br>damagin<br>g | PM1,PM2,PP2,PP3 |
| 25 | do | SLC34<br>A1 | KS8 | het | 2/4450288<br>9 | rs765538<br>592 | T>C | NM_00034<br>1 | V72A | NO | 1 | Probabl<br>y<br>damagin<br>g | PM1,PM2,PP3 |

**Table 2:**
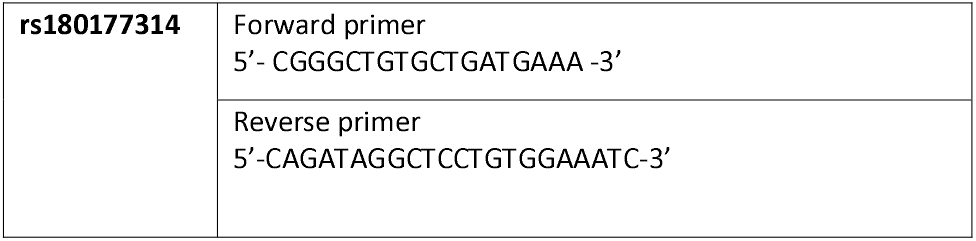
primers for sanger validation of rs180177314 of GRHPR gene.

When evaluating WES data for 3 known genes found in the cohort that cause hyperoxaluria was identified (Table 1), a single GRHPR mutation (rs180177314 (G>A) in 5 individuals (30%). Recessive or dominant causative mutations were detected in **19 gene**s (Table1). Pathogenic mutations were detected in 9 genes in among 18 individuals AGXT (3 individuals), ATP6V1B1 (1 individual), GRHPR (5 individuals,), HOGA1(2 individuals), FBN(1 individual), HSPG2 (1 individual), CRTAP(1 individual), VDR(1 individual), SLC34A1(1 individual).The family history, status of consanguinity, and detailed phenotype of individuals is shown **(supplementary table 1)**. The pedigrees of all homozygous mutations showed family history.

Among 18 individuals, within whom we detected causative mutations for hyperoxaluria, 9 detected mutations were novel pathogenic variants(Table 1), and **rs180177314 (G>A) in homozygous condition was reported and studied for the first time[BankIt2492453 seq2 MZ826703]** that have not been previously reported in databases of human disease-causing mutations. The detection rate of causative mutations was not different between sexes. The median age of onset was significantly lower in patients with a monogenic cause (most young) versus those without detection of monogenic cause (least young).We evaluated our cohort for differences regarding disease NL at presentation, age of onset of disease, and causative mutation detection. Moreover, a common snp rs141428607 G>T of SLC25A5 gene variant was found in 16 samples of the 18 studied patients. On the other hand, StringDB showed a direct interaction of SLC25A5 with GRHPR gene (fig6).

### Biochemical analysis

Among 18 paediatric KSD patients, 9were males and 9were females. There is a significant difference in patients and controls cohorts in terms of age (p value0.05). It is seen that blood urea and creatinine were much higher in patients having mutation in hyperoxaluria genes. Moreover urinary calcium excretion level and serum calcium level found to **higher** in patients (supplementary 2).

Furthermore, in this study we correlated the genotype to phenotype association for both the case and control study cohort in correspondence of genotype with respect to calcium level of both urine and serum, serum urea and serum creatinine. The rs180177314 (G>A) genotype shows a significantly in comparison to GG. whereas for 24-hour urinary calcium release also indicates a high level with genotype AA in contrast to GG.

### Characterization of kidney stones from samples

EDS analysis showed higher weight percentage of calcium 23.97 in GRHPR mutation with respect to normal kidney stone patients11.16 (Fig 2)

**Fig:2.**
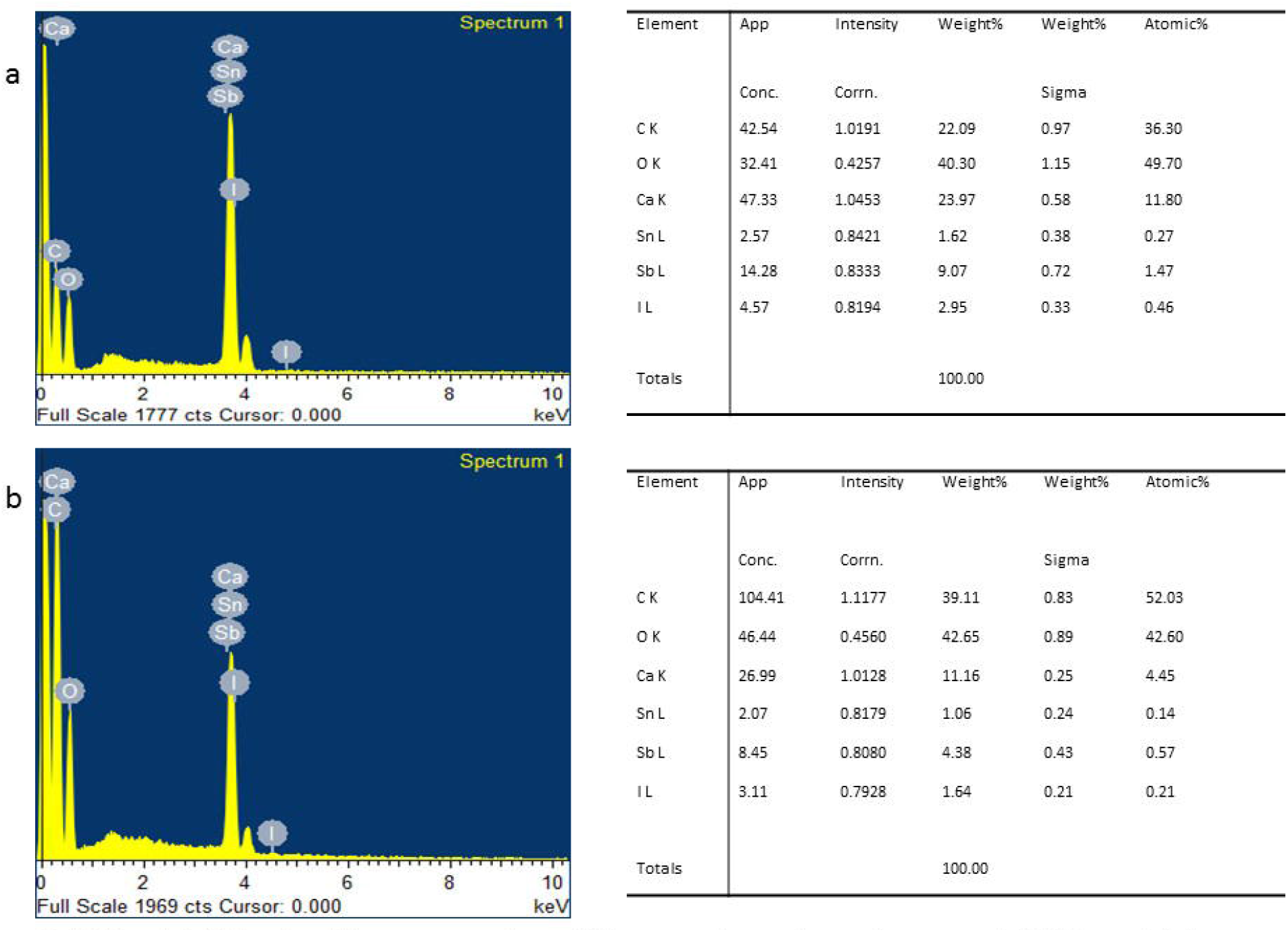
EDS analysis (b) showing calcium percentage of normal kidney stone where as increased percentage in GRHPR mutation(a)

**GRHPR G165D** rs180177314 (G>A

### R statistical analysis/ Association study

For variants (rs180177180), the G >A allele frequency distribution is significantly higher in the patient population (29.2%) compared to controls (5.9%). Our result indicates a 6.59-fold increased risk in disease progression with individuals carrying A allele In dominant model. Similarly, AA (29.2%) genotype is higher in case group and genotypic statistical codominant model (GG/GA+AA) confirm a increased risk in formation of stone. The genotype distribution for both of the SNPs are fitted the Hardy – Weinberg Equilibrium. Table 4

### Bioinformatics

Positive phylop scores show a value of 5.408, which predicts conservation in the genetic sequence and whose evolutionary change slower than expected, at locations that are likely to be conserved. On the other hand, phastcons value 0.832 explains that short highly-conserved regions and long moderately conserved regions can both obtain high scores. which in our case proves it. The mutant models showed high TM-score and low RMSD value in comparison with low TM-score and high RMSD value in the NaDH-binding domain(fig:3). A higher RMSD value indicates the greater structural dissimilarity between the wild type and mutant model. Consurf server predicted a high conversation site in G165D with a score of 9 with a confidence interval of -1.366,-1.117. The Project HOPE server revealed that the mutant residue is of bigger sizes, hydrophobic as well as negatively charge **(fig:4)** than the wild-type residue and these variations in size and hydrophobicity can disrupt the H-bond interactions with the adjacent molecules. Moreover, phyre2 it provides special backbone conformation but glycine replacement may interrupt that formation (The flexibility and rigidity of a protein structure are essential for exhibiting specific functions. Here, the analysis showed that amino acid substitution in G165D can disrupt flexibility and decreases stability. **Further docking results explain the Entropy-Enthalpy calculation for the analysis binding. the binding affinity protein-ligand molecular docking. _**

**Fig3:**
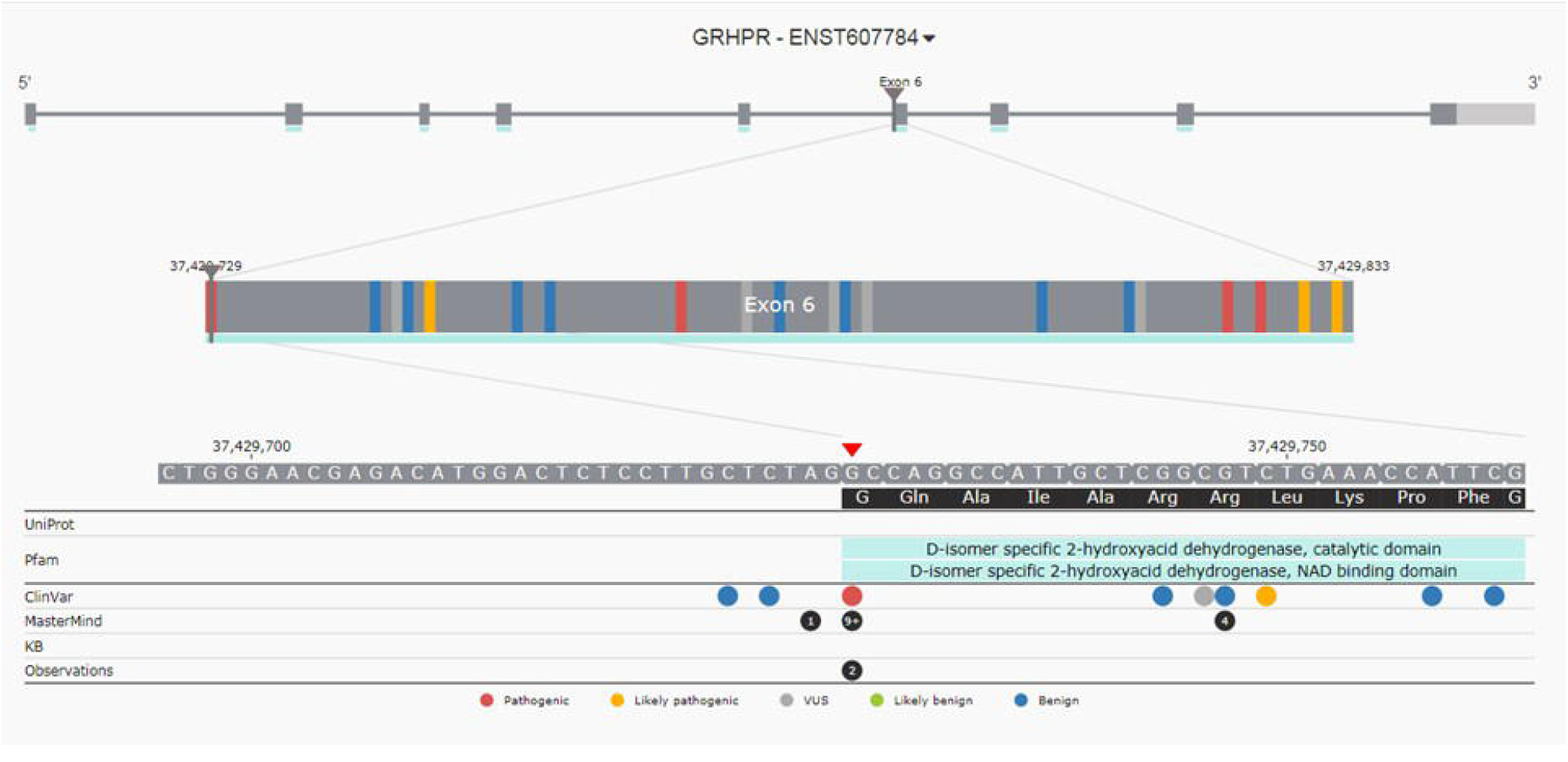
Figure showing mutation in the region of NaDH binding domain.

**Fig4:**
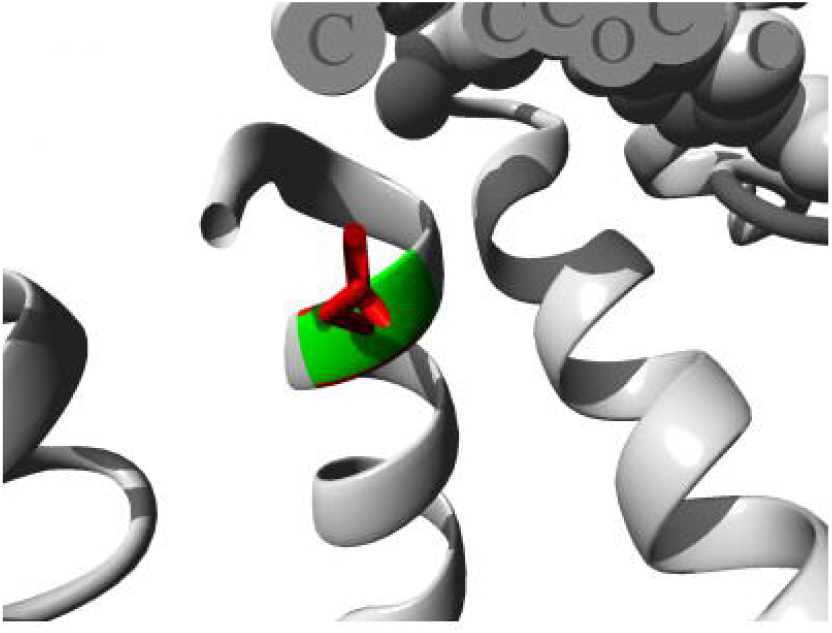
Mutated residue(ASP) shown in red and wild residue shown in green.

### RT-qPCR

The qRTpcr studies revealed that GRHPR is produced in human monocytes. Using the cDNA prepared from monocytes cells as mentioned above in the methodology. RT-qPCR was used to evaluate the expression of the GRHPR gene in human monocytes. In cases and controls cDNA was amplified using GRHPR gene-specific primers (Table 3) and SYBR Green to assess GRHPR gene expression. The expression of the GRHPR gene was shown to be greater in monocytes with the GRHPR mutation. [Fig2, compared to control samples] Furthermore, by examining the ΔΔCT value, RTqPCR data revealed that GRHPR gene expression was higher in case compared to control (fig 5).

**Table 3:**
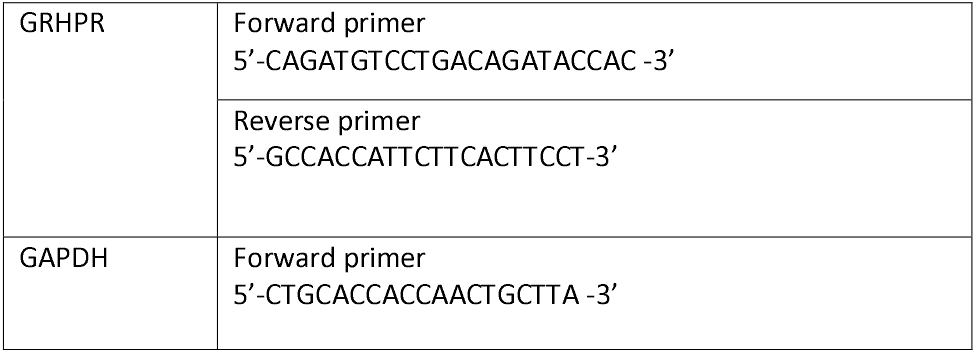
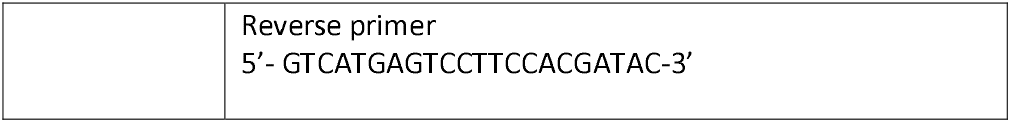
primer for RT_qPCR.

**Table 4:** Chi square test used to compare genotype frequencies between controls and patients

| SNP | <u>Genotype</u> | <u>Control (51)</u> | <u>Case (24)</u> | <u>MODEL</u> | <u>Odd ratio</u><br><u>(95% CI)</u><sup>a</sup> | <u>p-value</u> |
| --- | --- | --- | --- | --- | --- | --- |
| rs<br>**rs180177314** | GG | 48<br>(94.1%) | 17<br>(70.8%) | Codominant | GGVSGA<br>GGVS AA<br>0 | **0.0001** |
|  | GA | 3<br>(5.9 %) | 0<br>(20.0%) | Dominant | GG vs GA<br>+ AA<br>6.59<br>(1.53-28.4) | **0.007** |
|  | AA | 0<br>(0%) | 7<br>(29.2%) | Recessive | GG + GA<br>vs AA<br>0 | **0.00017** |

**Fig:5.**
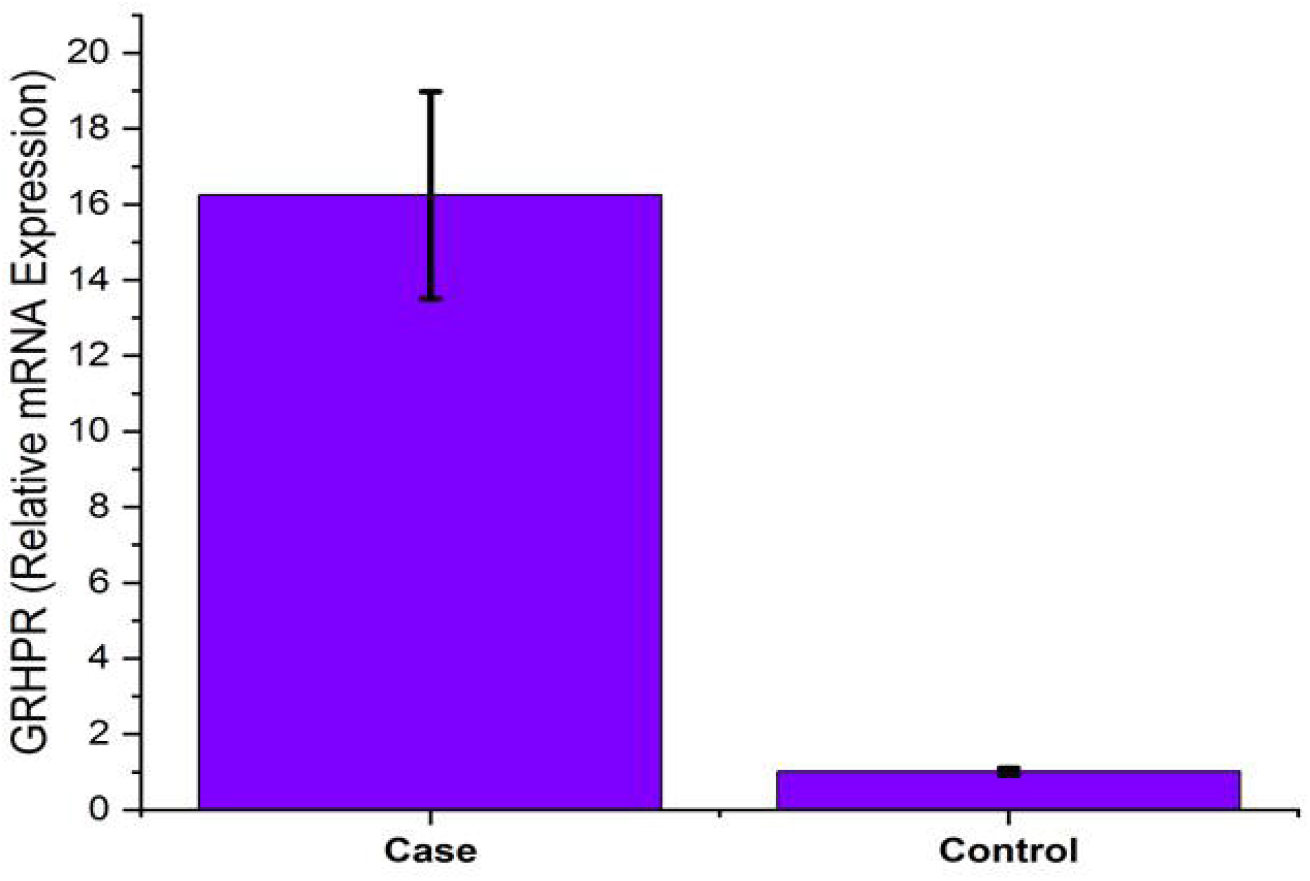
GRHPR gene expression status in monocytes.

**Fig6:**
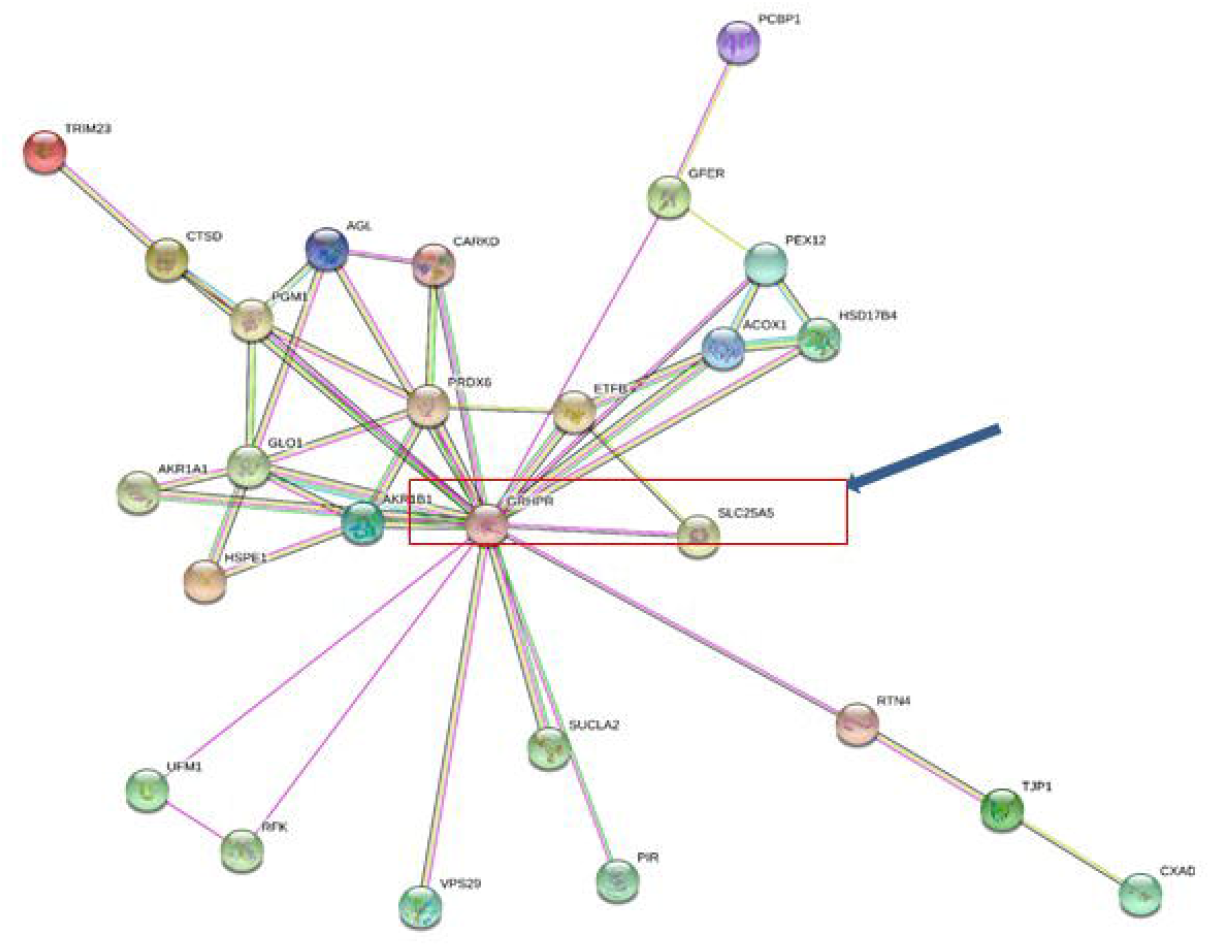
String analysis showing strong correlation between GRHPR and SLC25A5.

### Elisa

The results are shown as the mean SD of all rat samples (n = 25) at different days during the ethylene glycol therapy as described above. At different days of treatment, the fold change in protein expression between monocytes GRHPR levels and monocytes GAPDH levels revealed increased synthesis of GRHPR enzymes**(data not provided here)**

### Western blot

GRHPR signaling mediates the conversion of glyoxylate to glycolate. Mutation in the NAD binding domains alters the stability of the enzyme. We investigated the impact of GRHPR G165D homozygous mutation from patients samples described in methodology. In samples where mutation were present GRHPR upregulated (**fig7**) whereas in control expression was barely visible. indicating these proteins was upregulated to compensate for defective enzymes due to mutation.

**Fig7:**
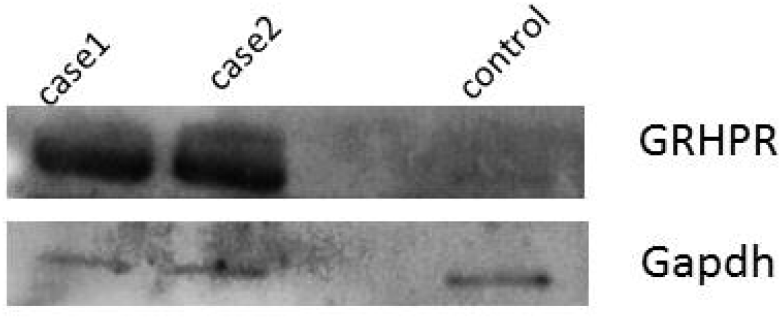
Shows monocytes expression of case samples (lane1 and lane2) and control monocytes expression in (lane4)

**Fig8:**
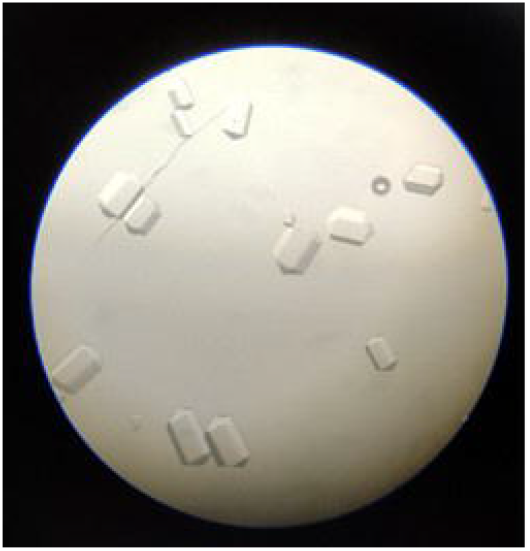
showing oxalate crystal in mouse urine.

## Discussion

Child nephrolithiasis is a less frequent disease. In this study, our surveyed data statistics showed that only 5-10% are children among population stratification of different age groups. Nephrolithiasis patients (reference with global data) our collected samples showed that a greater number of GRHPR and AGXT gene mutations among the studied age group to be more common.

Mutation in rs180177314 of GRHPR gene, PhyloP scores 5.08 can be concluded to determine how evolutionary conserved individual alignment sites. The evolution expected under neutral drift is contrasted to the interpretations of the scores. On the other hand PhastCons score of 0.832 is a hidden Markov model-based method that uses multiple alignment to estimate the chance that each nucleotide belongs to a conserved element. The phastCons values range from 0 to 1 and represent the probability of negative selection

The function of the enzyme, which is needed to maintain the cytosolic concentration of hydroxypyruvate and glyoxylate at a very low level, thus preventing the formation of oxalate GRHPR, present in excess because of deficiency in the enzyme that converts it to D-glycerate, stimulates oxidation of glycolate to oxalate, and decreases reduction of glyoxylate to glycolate. This is a novel explanation for the phenotypic consequences of a garrodian inborn error of metabolism. D-glycerate dehydrogenase also has glyoxylate reductase activity. The 2 activities are attributable to a single enzyme. The deficiency of D-glycerate dehydrogenase activity presumably causes accumulation of its substrate, hydroxypyruvate, which is then converted to L-glycerate by the action of L-lactate dehydrogenase. **The deficiency of glyoxylate reductase activity <u>presumably</u> causes the impaired conversion of glyoxylate to glycolate**. Conversion of glyoxylate to oxalate by L-lactate dehydrogenase would explain the observed hyperoxaluria. As in type I primary hyperoxaluria, the main clinical manifestation is calcium oxalate nephrolithiasis.

Clinically, a simple activity test can reveal the causative mutation in paediatric nephrolithiasis. Also AGXT, GRHPR and HOGA1 are intrerlinked through various pathways. The underlying cause of SLC25A5 with GRHPR is yet to be studied in relation to renal stones and SLC25A5 has a role in the calcium homeostasis pathway but its relation to hyperoxaluria is still to be studied.

## Conclusion

We have studied the underlying cause of kidney stoney in lower age group. WES revealed that most lower age group patients are prone to hyperoxaluria. Patient’s monocytes from blood samples were used for expression and activity analysis. Data obtained from the western blot and RT-qPCR showed increased production of GRHPR enzymes with respect to normal samples and other KSD patients. but the activity of the enzyme was negligible. When other chemically induced hyperoxaluria was carried out in the RAT model it also showed increased production as well as the activity of the enzyme. Hence, GRHPR c.494G>A is a <u>marker</u> and causes child nephrolithiasis in the e**astern part of India**.

## Data Availability

All data produced in the present work are contained in the manuscript

## Authors Declaration

**Authors have no conflict of interest**.

