## supplementary tables for "Whole Exome Sequencing (WES) identifies Homozygous mutation in g.37429732G>A in GRHPR gene is responsible for early onset of nephrolithiasis in the population of West Bengal, India"

| Sample ID | Gender | Onset of Disease | Kidney stone sample | Familial history(M,F,S)  Present =1  Absent=0  No data=? | consanguinity | Self history of kidney stone | **Clinical phenotype**  **Other than KSD** |
| --- | --- | --- | --- | --- | --- | --- | --- |
| KS1 | Male | 8 | present | M0, F0 | no | Yes | hyperoxaluria |
| KS2 | Female | 6 | present | M0, F? | no | No | Not found |
| KS3 | male | 2 | present | M0, F? | no | No | Renal tubular disfunction |
| KS4 | female | 8 | present | M0, F0 | no | Yes | hyperoxaluria |
| KS5 | female | 11 | present | M1, F? | No | No | Not found |
| KS6 | female | 11 | present | M1, F? | No | yes | hyperoxaluria |
| KS7 | male | 13 | present | M1, F0 | No | yes | Not found |
| S4 | female | 10 | present | M0, F0 | No | yes | hyperoxaluria |
| S13 | female | 14 | present | Present m??, F?? | No | yes | hyperoxaluria |
| KS8 | male | 6 | present | M? ,F? | No | No | No data available |
| KS9 | male | 2 | present | 0 | No | No | No data available |
| KS10 | male | 2 | present | Present, M?, F? | No | No | hyperoxaluria |
| KS11 | female | 3 | present | ? | No | No | Not found |
| KS12 | male | 9 | present | M0,F0 | No | yes | CKD, Hyperoxaluria |
| KS13 | male | 11 | present | F1 | No | yes | Not found |
| KS14 | female | 6 | present | M0, F0 | No | No | Hyperoxaluria, Hydroneprosis |
| KS15 | Male | 9 | present |  | No | No | Hydronephrosis |
| KS16 | male | 10 | present | M0, F0 | No | yes | Not found. |

Supplementary table 1- Details of patient sample

| Sl no | Sample id | Gender(*) | mutation | previous history of stones(*) | Blood urea | Creatinine | Serum Calcium | 24 hr urine sample calcium/ phosphate (mg/day) | Diameter of stone (cm) |
| --- | --- | --- | --- | --- | --- | --- | --- | --- | --- |
| 1 | KS1 | M | grhpr | yes | 36 | 1.7 | 9.5 | 45/167 | 3.5 |
| 2 | KS4 | F | agxt | yes | 13 | 0.9 | 10 | 27/167 | 1.2 |
| 3 | KS6 | F | agxt, Hoga1 | yes | 60 | 6.5 | 12 | 37/165 | 4 |
| 4 | KS10 | M | grhpr | no |  |  |  | 41/150 | 3.2 |
| 5 | KS12 | M | grhpr | yes | 163 | 6.6 | 7.6 | 77/162 | 4.2 |
| 6 | KS14 | F | grhpr | yes | 45.25 | 3.65 |  | 40/155 | 3.7 |
| 7 | S4 | F | grhpr, agxt | yes | 60 | 2.5 | 10.5 | 67.5/145 | 6 |
| 8 | S13 | F | hoga | yes | 13 | 0.8 | 9.5 | 57/215.6 | 4.2 |

Supplementary table 3: Biochecmical data.
